# Patient experience with healthcare: Feedback for a ‘Post COVID-19 clinic’ at a tertiary care center in rural area

**DOI:** 10.1101/2021.11.20.21266640

**Authors:** Alpana Garg, Maran Subramain, Patrick B Barlow, Lauren Garvin, Karin F Hoth, Kimberly Dukes, Richard M Hoffman, Alejandro P. Comellas

**Affiliations:** Department of Internal Medicine, Division of General Internal Medicine, University of Iowa, IA, Iowa; Institute for Clinical and Translational Science, University of Iowa, IA, Iowa; Department of Psychiatry, University of Iowa, IA, Iowa; Iowa Neuroscience Institute, University of Iowa, IA, Iowa; Center for Access & Delivery Research and Evaluation, Iowa City VA Healthcare system; Holden Comprehensive Cancer Center, University of Iowa, IA, Iowa; Department of Internal Medicine, Division of Pulmonary, Critical Care and Occupational Medicine, University of Iowa, IA, Iowa

**Keywords:** Post-Acute Sequalae of SARS-CoV-2 infection (PASC), post-acute COVID-19 syndrome, Post-Covid-19 Clinic, rural health, patient experience, Long-COVID

## Abstract

**Purpose:** Post-acute sequelae of SARS-CoV-2 (PASC) is a complex condition with multisystem involvement. We assessed patients’ perspectives and experience with a PASC clinic established at University of Iowa in June 2020.

**Methods:** We conducted a mixed-method survey in June 2021 to ask PASC clinic patients about 1) PASC symptoms and their impact on physical and mental health, and cognition using the PROMIS Global Health and Cognitive Function abilities items, and 2) satisfaction with clinic services and referrals, barriers to care, and recommended support resources.

**Findings:** Ninety-seven patients (97/277, 35% response rate) completed the survey. Most were women (67%, n=65/97), Caucasian (93%, n=90/97) and received outpatient care during acute COVID-19 illness (79%). Fifty percent reported wait time of 1-3 months and 40% traveled >1 hour for appointment. The most common symptoms >3 months from initial infection were fatigue (77%), “brain fog” (73%), exercise intolerance (73%), anxiety (63%), sleep difficulties (56%) and depression (44%). Qualitative analysis of open-ended answers added valuable context to quantitative results. A minority of patients reported significantly reduced functioning (≥1.5 SD below mean) of their physical health (22.5%), mental health (15.9%) and cognitive abilities (17.6%). Satisfaction with clinical services was high though participants identified barriers to care including scheduling delays and financial concerns. Respondents suggested potential strategies for optimizing recovery including continuity of care, a co-located multispecialty clinic and being provided with timely information from emerging research.

**Conclusion:** Our study reports high PASC symptom burden, its impact on health and patient experience with healthcare. It is important that primary healthcare professionals listen to patients with empathy and support them during recovery. Healthcare systems and policymakers should focus on accessible, comprehensive, and patient-centered integrated care.

## Introduction

Post-acute sequelae of SARS-CoV-2 (PASC) is a complex condition with multisystem involvement.^1-6^ In a longitudinal study 87.4% patients who recovered from COVID-19 had at least one symptom at 60 days of follow-up.^7^ At 9 months’ follow-up, studies report the presence of one or more symptoms in 30-36% of survey respondents.^8, 9^ A recent report of 342 patients, showed that 40.7% had at least one symptom at one year of follow-up.^10^ The persistence of symptoms can significantly affect patients’ quality of life.^2, 7, 11, 12^

Rural areas were disproportionately affected by SARS CoV-2 infection and people living in underserved communities face unique barriers to optimal care.^13-17^ Currently in Iowa, more than 500,000 cases and 7200 deaths have been reported with major outbreaks affecting the diverse work force in meat packaging plants in Iowa, United States (US).^18, 19^ Mortality rate in some rural counties in Iowa (290 deaths per 100,000) has surpassed the national US mortality rate (200-225 deaths per 100,000).^18^ University of Iowa Hospitals and Clinics (UIHC) is one of the largest academic tertiary healthcare systems in this area and caters to the healthcare needs of Iowa and neighboring states.

Healthcare institutions must adapt to the needs of growing patient population with PASC and there is a need to understand patient’s perspectives to develop an optimally functioning specialty clinic to provide appropriate care to assist in patient recovery. Accordingly, we surveyed patients who attended our PASC clinic about their perspectives and experiences.

## METHODS

### Structure of the PASC Clinic

The clinic is staffed by nine providers (including intensivists and internists). Patients are eligible to be seen 30 days after a documented positive SARS-CoV-2 test (either positive nasopharyngeal RT-PCR or antibody test). The clinic operates one full day per week, and cares for an average of 15-20 patients per day in 40-minute appointment slots. Patients are evaluated either by a pulmonary specialist or an internal medicine physician. New patients are triaged by a nurse coordinator for appropriateness of lung testing (not ordered if no respiratory complaints or tested already elsewhere). However, during their first appointment, most patients undergo a pulmonary function test, computed tomography (CT) of the chest, laboratory testing (complete blood count and comprehensive metabolic panel), and screening for depression (Patient Health Questionnaire-9) and anxiety (Generalized Anxiety Disorder-7).^23, 24^ Patients are assessed for dyspnea using the mMRC (Modified Medical Research Council) dyspnea scale, documenting all symptoms, and discussing test results with the patient.^25^ Patients are referred for subspecialty evaluation as appropriate.

This study received Institutional Review Board approval (#202105502). Patients who attended PASC clinic at UIHC through the in first year after clinic inception (June 12^th^ 2020), were invited to complete an online mixed-methods survey. By completing the survey, patients provided consent to participate. Patients were emailed information about the study and a link to the survey, with two weekly reminders sent to non-respondents.

### Instruments

The survey (Appendix 1) was drafted using Qualtrics (Qualtrics, Provo, UT). We collected demographic data (age, sex, and ethnicity) and asked about the 1) timing of initial SARS-CoV-2 infection, highest level of care utilized, symptoms at the time of initial infection, symptoms three months post-infection and up to four most concerning symptoms at the time of the survey; 2) impact of symptoms on perceived physical and mental health, assessed by the Patient Reported Outcomes Measurement Information System Global Health-10 item questionnaire (PROMIS GH-10) and effect on cognitive ability assessed by the PROMIS Cognitive Function-Abilities 4a-4 item questionnaire (PROMIS CA-4a);^26-28^ and 3) experiences with the PASC clinic including wait time for first appointment, distance traveled, satisfaction with services (scheduling, testing, imaging, staff), subspecialty referrals provided and completed, barriers to attending subspeciality referrals, and recommended resources for other patients with PASC. Participants also answered open-ended questions, identifying any other resources that would be helpful in an “ideal post-COVID clinic” and adding any other comments.

### Data Analysis

Non-responder analysis was run to compare demographics and key clinical characteristics between survey responders and non-responders using chi-square tests of independence and independent sample t-tests as appropriate. Differences were expressed in terms of mean differences or odds ratios with 95% confidence intervals (CI).

Symptoms during initial infection and at three months post-infection were compared within respondents using a sign test to determine the shift in a respondent’s symptoms over time; “positive shift” indicated the number of symptoms increased over time, a “negative shift” indicated symptoms decreased, and a “tie” indicated that the number of symptoms stayed the same. Internal consistency and reliability of the two PROMIS instruments in this sample was established using a Cronbach’s alpha with α = 0.60 being sufficient and α ≥ 0.80 being ideal. Once confirmed, raw scores for the physical and mental health components of the PROMIS GH-10 and the PROMIS CA-4a were transformed into normatively referenced t-scores with a mean of 50 and SD of 10 per scoring guides.^28, 29^ These scores were classified as either “normal” (or within normal limits) defined as mean ±1.49 SDs or as “significantly reduced” at ≥1.5 SDs below the mean.^30, 31^ Sex and type of care received during initial infection were used to compare the odds of scoring within normal range or reduced range on the PROMIS measures using chi-square tests of independence and Fisher’s exact tests, as appropriate. The Mann-Whitney U test was used to compare the number of symptoms at initial infection and at three-months post-infection in respondents whose PROMIS scores were reduced vs. those in the normal range. All statistical tests were conducted using IBM SPSS Statistics v.27 (IBM, inc. Armonk, NY), were two-sided, and p ≤0.05 was considered statistically significant. Thematic analysis on the open-ended responses was conducted to identify general categories of information.^32^

## RESULTS

### Demographics

The survey response rate was 35% (97/277). Most respondents were women (67%), Caucasian (92.8%) and were diagnosed with SARS-CoV-2 between March 2020 and April 2021. Non-Caucasian patients were over three times more likely to be non-responders compared to Caucasian patients, with an odds ratio (OR) of 3.06 (95% CI = 1.23-7.64, *p* = 0.013).

### Level of Care Received During Initial COVID-19 Infection

All respondents were at least 64 days from their initial positive test when they completed the survey, with a median interval of 234 days (IQR = 124). Overall, 74 (79%) of respondents reported receiving outpatient care including self-management at home, telemedicine visits only, urgent care visits, and/or outpatient emergency room visits. Comparatively, 23 respondents (24%) reported being admitted either to the hospital or ICU, or both (a respondents often had multiple levels of care; thus, percentages do not equal 100%). Men (75%) and women (81.5%) were similarly likely to receive care in an outpatient or home setting, (*p* = 0.45). Women were less likely to be admitted to the hospital or ICU (22%) than men (28%), which was not statistically significant (*p* = 0.47).

### Characteristics of Initial SARS-CoV-2 infection and PASC illness

Symptoms were classified into six broad body systems: general, neurological/nerves/memory, lung, heart, gastrointestinal, and mental health (Appendix 1). At the time of diagnosis, the most common symptoms reported were fatigue (88%), shortness of breath (73%), exercise intolerance (73%) and palpitations or heart racing (73%). The most common reported symptoms three months post-infection included fatigue (77%), “brain fog” or memory or cognitive impairment (73%), exercise intolerance (73%) and shortness of breath (70%; Figure 1).

**Figure 1.**
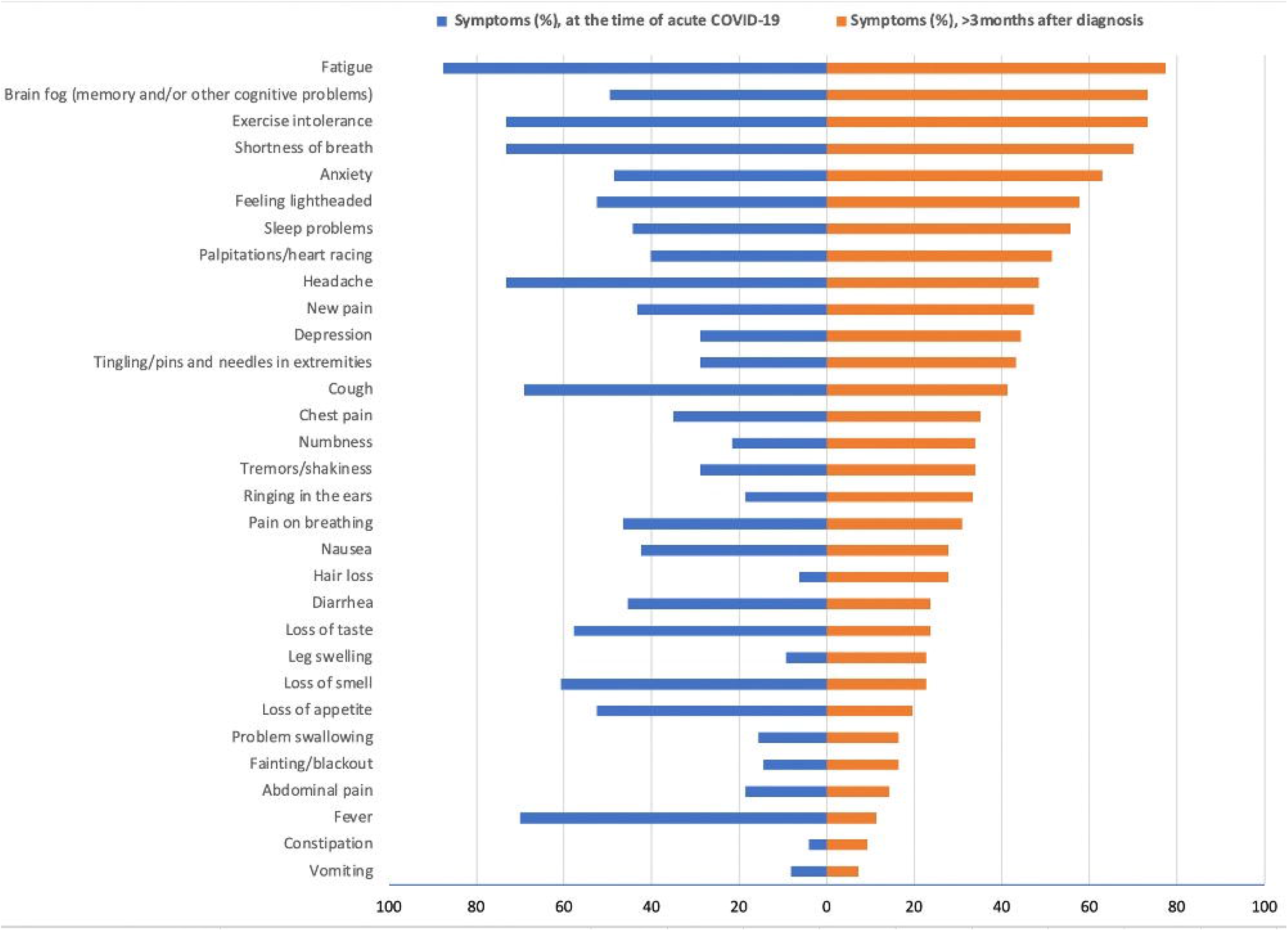
Symptoms at time of diagnosis and more than three months after SARS-CoV-2 infection.

Respondents were asked to report most concerning symptoms that they were experiencing at the time of survey (Table 2). Fatigue (52%), “brain fog” (39%), and shortness of breath (30%) were the most cited ongoing concerns, followed by cough (22%). Women generally reported a higher prevalence of all symptoms compared to men (as shown in Table 2), particularly fatigue (15 percentage points more) and cough (26.2 percentage points more), but far fewer men than women provided answers to this open-ended question.

**Table 1.** Demographic comparisons between responders and non-responders.

| Variable, group | Response group mean (SD) or frequency (%) |  | Test statistic (95% CI) | *p* value<sup>2</sup> |
| --- | --- | --- | --- | --- |
|  | Yes (*n* = 97) | No (*n* = 180) |  |  |
| Age | 48.66 (14.77) | 44.81 (15.84) | MD* = 3.85 (0.01, 7.69) | 0.049 |
| Sex |  |  |  |  |
| Male | 32 (36.4%) | 56 (63.6%) | OR = 1.09 (0.64, 1.85) | 0.749 |
| Female | 65 (34.4%) | 124 (65.6%) |  |  |
| Race/Ethnicity† |  |  |  |  |
| White | 90 (38.0%) | 147 (62.0%) | OR = 3.06 (1.23, 7.64) | 0.013 |
| Non-White | 6 (16.7%) | 30 (83.3%) |  |  |

MD, mean difference, OR odds ratio
\*Mean difference analyzed with independent *t*-test, and frequencies compared using chi-squared test of independence.
†Groups were combined due to low and disparate sample sizes. The largest racial/ethnicity identity aside from White was Latino/Latinx (*n* = 22).

**Table 2.** Patients’ self-reported most important current PASC symptoms (at median of 234 days after diagnosis of COVID-19).

| Current symptom | Frequency (%) * |  |  |
| --- | --- | --- | --- |
|  | Male | Female | All participants |
| Fatigue | 8 (42.1%) | 20 (57.1%) | 28 (51.9%) |
| Brain fog | 7 (36.8%) | 14 (40%) | 21 (38.9%) |
| Shortness of breath | 5 (26.3%) | 11 (31.4%) | 16 (29.6%) |
| Cough | 1 (5.3%) | 11 (31.4%) | 12 (22.2%) |
| Exercise intolerance | 1 (5.3%) | 6 (17.1%) | 7 (13%) |
| Anxiety | 1 (5.3%) | 1 (2.9%) | 2 (3.7%) |

\*Participants could enter up to four different symptoms, so total percent exceeds 100%; percentages were calculated out of the total number of participants who responded to these questions.

The median number of general symptoms reported was 3.0 for both the initial infection and three-month post-infection periods; however, 42.3% of participants reported fewer symptoms at three months compared to baseline, 20.6% reported more symptoms, and 37.1% reported the same number of symptoms (p = 0.01; Table 3). Only 15.5% of patients reported decreased symptoms at three months compared to 29.9% reporting increased symptoms (*p* = 0.05).

**Table 3.**
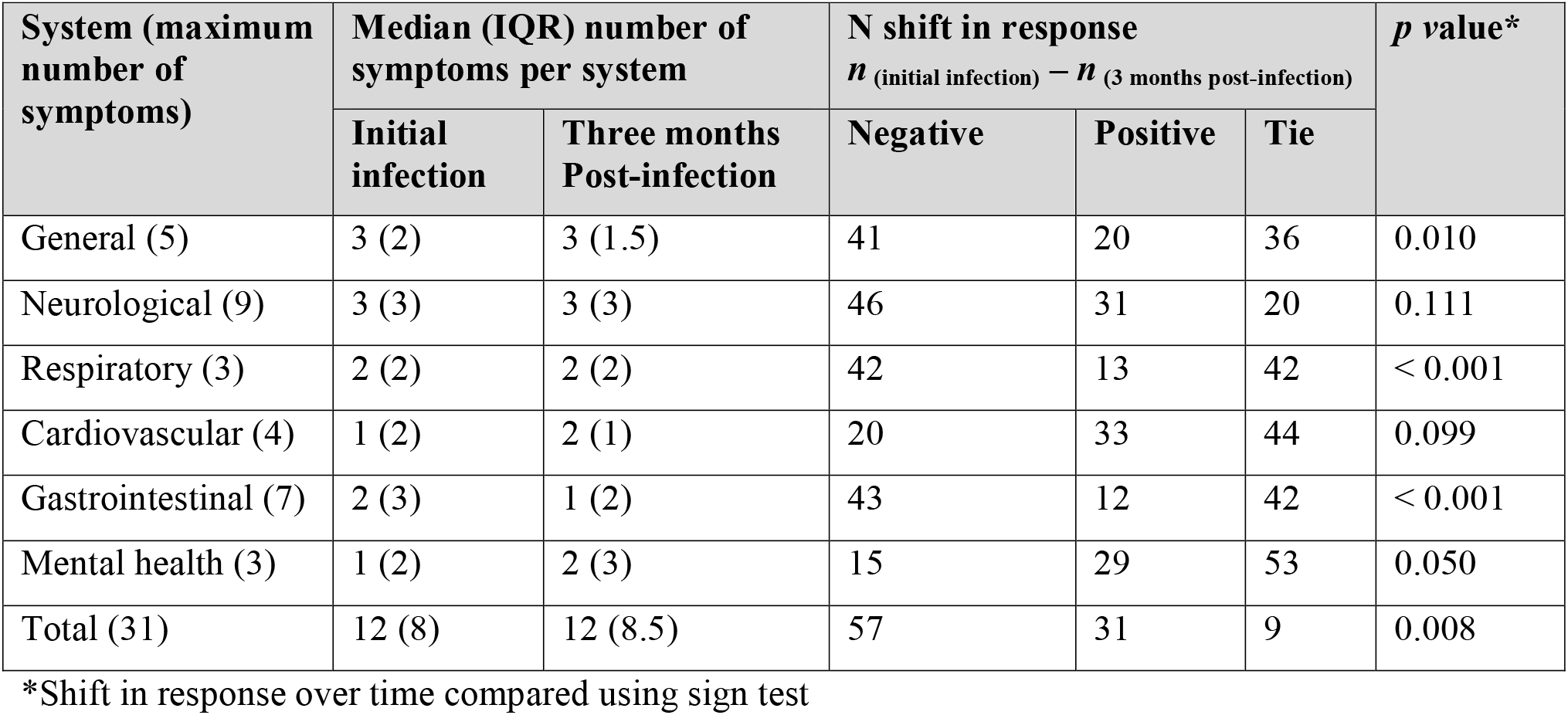
Change in total symptom frequency over time.

| System (maximum number of symptoms) | Median (IQR) number of symptoms per system | | N shift in response<br>$n$ (initial infection) – $n$ (3 months post-infection) | | | *p* value* |
| --- | --- | --- | --- | --- | --- | --- |
|  | Initial infection | Three months Post-infection | Negative | Positive | Tie |  |
| General (5) | 3 (2) | 3 (1.5) | 41 | 20 | 36 | 0.010 |
| Neurological (9) | 3 (3) | 3 (3) | 46 | 31 | 20 | 0.111 |
| Respiratory (3) | 2 (2) | 2 (2) | 42 | 13 | 42 | < 0.001 |
| Cardiovascular (4) | 1 (2) | 2 (1) | 20 | 33 | 44 | 0.099 |
| Gastrointestinal (7) | 2 (3) | 1 (2) | 43 | 12 | 42 | < 0.001 |
| Mental health (3) | 1 (2) | 2 (3) | 15 | 29 | 53 | 0.050 |
| Total (31) | 12 (8) | 12 (8.5) | 57 | 31 | 9 | 0.008 |

\*Shift in response over time compared using sign test

### Impact on Physical and Mental Health, and Cognition

The total Cronbach’s alpha coefficients for the PROMIS GH-10 and PROMIS CA-4a were α = 0.78 and α = 0.96, respectively; thus, both instruments were considered acceptably reliable for our sample. The distribution of scores for PROMIS GH-10 and PROMIS CA-4a are shown in Table 4. The odds of expressing reduced physical health, mental health, and cognitive function did not differ by sex or initial care (outpatient vs. inpatient; Appendix 2). In contrast, we found significant differences between patients expressing normal (mean ±1.49 SD) versus significantly reduced (≥1.5 SD) physical health, mental health, and cognitive abilities across five of six body systems as well as median total symptom burden at three months post-infection (Table 5). Those showing reduced mental health reported double the average neurological symptoms (6, IQR = 2) compared to those who did not report reduced mental health (3, IQR = 2.25, *p* < 0.001). Similarly, the total number of symptoms reported by those with reduced mental health was 7.5 greater (median = 18.5, IQR = 5.25) than participants without (median = 11, IQR = 6.5, *p* < 0.001).

**Table 4.** Distribution of PROMIS GH-10 physical and mental health sub-scores and the PROMIS Cognitive Function Abilities 4a on t-distribution (M = 50, SD = 10). Only participants with responses to all items are included in the calculations.

| *t*-Score | PROMIS GH-10 ($\alpha = 0.78$) | | | | Cognitive function abilities ($\alpha = 0.96$) | |
| --- | --- | --- | --- | --- | --- | --- |
|  | Global physical health |  | Global mental health |  |  |  |
|  | *n* | Valid | *n* | Valid | *n* | Valid |
| $\geq 1.5$ SDs below mean | 17 | 22.7% | 14 | 15.9% | 16 | 17.6% |
| Between 1.49 SDs below mean and mean | 44 | 58.7% | 62 | 70.5% | 53 | 58.2% |
| Between mean and 1.49 SDs above mean | 14 | 18.7% | 12 | 13.6% | 22 | 24.2% |
| $\geq 1.5$ SDs above mean | 0 | 0 | 0 | 0 | 0 | 0 |

**Table 5.** Median number of symptoms experienced at baseline and three months post-infection by score group on PROMIS measures.

| Time period and system<br>(maximum number of<br>symptoms) | Physical health group median (IQR) |  | *p* value* |
| --- | --- | --- | --- |
| | Normal | Reduced $\geq 1.5$ SD | |
| Initial Infection |  |  |  |
| General (5) | 3 (2) | 3 (1.5) | 0.713 |
| Neurological (9) | 3.5 (2) | 4 (3) | 0.217 |
| Respiratory (3) | 2 (2) | 2 (2) | 0.680 |
| Cardiovascular (4) | 1 (2.25) | 2 (1) | 0.825 |
| Gastrointestinal (7) | 2 (3) | 3 (3) | 0.651 |
| Mental health (3) | 1 (2.25) | 1 (3) | 0.469 |
| Total (31) | 13 (8) | 14 (10) | 0.630 |
| Three months post-infection |  |  |  |
| General (5) | 3 (1.25) | 3 (1) | 0.004 |
| Neurological (9) | 3 (3) | 5 (2.5) | 0.000 |
| Respiratory (3) | 2 (1) | 2 (1.5) | 0.417 |
| Cardiovascular (4) | 2 (1) | 2 (1) | 0.027 |
| Gastrointestinal (7) | 1 (2) | 2 (2) | 0.039 |
| Mental health (3) | 2 (3) | 3 (1) | 0.006 |
| Total (31) | 11 (7.25) | 16 (5) | <0.001 |
|  | Mental health group median (IQR) |  | *p* value |
| | Normal | Reduced $\geq 1.5$ SD | |
| Initial infection |  |  |  |
| General (5) | 3 (2) | 3 (2) | 0.840 |
| Neurological (9) | 4 (3) | 3 (2.25) | 0.287 |
| Respiratory (3) | 2 (2) | 3 (1.25) | 0.078 |
| Cardiovascular (4) | 1 (2) | 1.5 (2) | 0.397 |
| Gastrointestinal (7) | 2 (3) | 2 (3) | 0.893 |
| Mental health (3) | 1 (2.25) | 0 (3) | 0.456 |
| Total (31) | 13 (8) | 12.5 (7.75) | 0.991 |
| Three months post-infection |  |  |  |
| General (5) | 2 (2) | 3 (1) | 0.004 |
| Neurological (9) | 3 (2.25) | 6 (2) | <0.001 |
| Respiratory (3) | 1.5 (2) | 2 (0.75) | 0.166 |
| Cardiovascular (4) | 2 (1) | 3 (1) | 0.001 |
| Gastrointestinal (7) | 0 (1.25) | 2 (2) | 0.006 |
| Mental Health (3) | 2 (3) | 3 (0.25) | <0.001 |
| Total (31) | 11 (6.5) | 18.5 (5.25) | <0.001 |
|  | **Cognitive function abilities group median (IQR)** |  | ***p* value** |
|  | **Normal** | **Reduced $\geq 1.5$ SD** |  |
| **Initial infection** |  |  |  |
| General (5) | 3 (2) | 3 (0.75) | 0.867 |
| Neurological (9) | 3 (2) | 5 (3.75) | 0.126 |
| Respiratory (3) | 2 (2) | 3 (1.75) | 0.082 |
| Cardiovascular (4) | 1 (2) | 1.5 (1) | 0.491 |
| Gastrointestinal (7) | 2 (3) | 3 (3.5) | 0.190 |
| Mental Health (3) | 1 (2) | 1 (3) | 0.948 |
| Total (31) | 12 (8) | 15 (9.5) | 0.122 |
| **Three months post-infection** |  |  |  |
| General (5) | 2 (2) | 3 (1) | <0.001 |
| Neurological (9) | 3 (2) | 5 (2.5) | <0.001 |
| Respiratory (3) | 2 (1) | 2 (2.75) | 0.662 |
| Cardiovascular (4) | 2 (1) | 2 (1) | 0.018 |
| Gastrointestinal (7) | 0 (2) | 1 (2) | 0.017 |
| Mental Health (3) | 2 (3) | 3 (1) | 0.003 |
| Total (31) | 11 (7) | 16 (4.75) | <0.001 |

\*All groups compared using Mann-Whitney U test

### PASC Clinic Care Wait and Travel Time

Fifty percent of patients reported waiting 1-3 months, whereas 37% were seen within one month of contacting the clinic for a first appointment. Only 8 patients reported waiting 3-4 months for their first appointment. Most (60%) patients traveled less than one hour for their clinic appointment.

### Referrals

Over 50% of our participants reported receiving one to three different specialty referrals after COVID-19 diagnosis, most commonly to pulmonology (50%), cardiology (27%), pulmonary rehabilitation (20%) and physical & occupational therapy (18%). (Figure 2) As symptom burden increased, the number of referrals to various subspecialists increased. There were moderate, positive correlations between total initial symptoms and referrals made (r = 0.45, *p* < 0.001) and total initial symptoms and referrals completed (r = 0.44, *p* = < 0.001).

**Figure 2.**
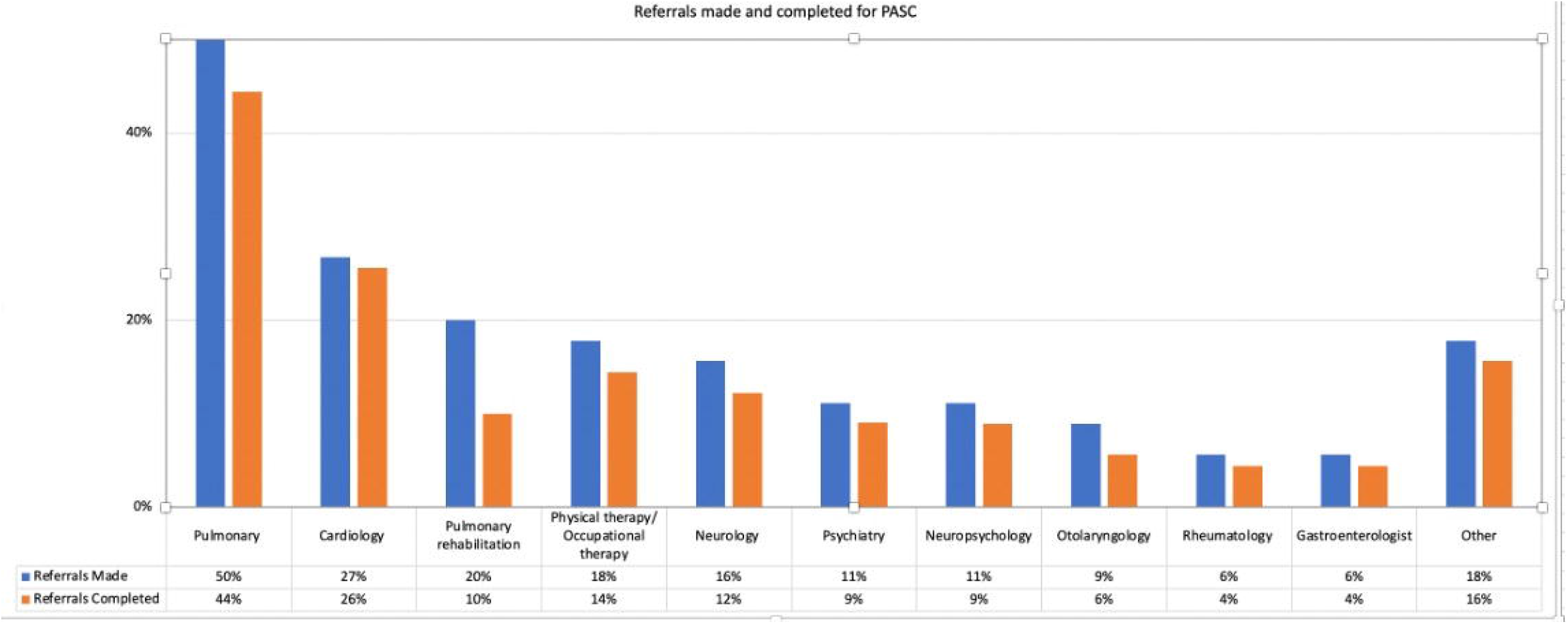
Referrals to specialists recommended and completed after SARS-CoV-2 infection. Other included open-ended responses like referral for sleep study, hematology-oncology, acupuncture and ‘no referrals were made’

### Barriers to referrals

On direct survey questions, the most common barriers reported for completing referrals were delays in getting appointments or not getting an appointment (21%) and financial concerns with uncertainty about insurance coverage for tests and appointments (16%). Other barriers reported included multiple clinic visits with different providers adding complexity of care (9%), travelling a long distance for the appointments (8%), inability to miss work to attend appointments (3%), limitations from symptom burden (3%), and preference for a virtual appointment rather than in-person due COVID-19 infection with in-person care (3%). Other barriers (18%) included responses such as “*waiting to see if symptoms get better*”, “*not sure which specialty to see*”, and “*no referrals were made*”, and “*insurance will not cover tests/physical therapy*”.

### Satisfaction with PASC clinic

Most participants (between 61% and 93.2%) indicated they were either “satisfied” or “very satisfied” with their experiences and the dissatisfaction rate with all the services was less than 20%. (Table 6)

**Table 6.** Participant satisfaction with PASC clinic experience. Values calculated from complete surveys only.

| How satisfied are you with your experience in the following categories? | Frequency (%) |  |
| --- | --- | --- |
|  | Dissatisfied or very dissatisfied | Satisfied or very satisfied |
| Scheduling clinic appointments | 10 (11.6) | 60 (69.8) |
| Getting lung function tests (also known as PFTs or spirometry) | 4 (4.7) | 73 (85.9) |
| Getting CT scan | 3 (3.8) | 67 (84.8) |
| Getting blood tests | 5 (7) | 54 (76.1) |
| Interaction with the front desk staff and medical assistant | 1 (1.1) | 82 (93.2) |
| Interaction with the doctor in the post-COVID clinic | 9 (10.2) | 71 (80.7) |
| Scheduling referrals / care coordination for subspecialties like cardiology, psychiatry, rheumatology, rehabilitation medicine (OT or PT) | 6 (10.2) | 39 (66.1) |
| Overall, addressing your physical health concerns | 17 (19.8) | 55 (64) |
| Overall, addressing your mental health | 11 (14.3) | 47 (61) |

### Resource needs

Participants were asked to rate how helpful potential candidate resources and services might be in addressing PASC (Table 7). “Very helpful” or extremely helpful options included: 1) continuity of care i.e., a regular follow-up appointment scheduled with the same doctor (75%), 2) providing a multi-specialty clinic i.e., different doctors located in the same clinic space (70.4%), and 3) additional patient education from reliable sources (68.7%). Conversely, 30% or more found providing access to social workers, access to a scheduling coordinator, and group therapy interventions to be either “slightly helpful” or “not helpful at all.”

**Table 7.** Perceived helpfulness of future resources.

| How helpful do you think each of the following resources could be to you or other patients with post-COVID symptoms? | Frequency (%) |  |
| --- | --- | --- |
|  | Not or slightly helpful | Very or extremely helpful |
| Continuity of care and regular follow-up with same doctor(s) | 8 (9.5) | 63 (75) |
| Physical therapy/occupational therapy (rehabilitation to increase physical strength and exercise capacity) | 14 (17.3) | 50 (61.7) |
| Pulmonary rehabilitation to improve breathing (supervised program) | 13 (16) | 55 (67.9) |
| Informal support group open forum e.g., online support group with other patients with similar experiences | 18 (22.2) | 44 (54.3) |
| Group therapy intervention led by a mental health provider (e.g., online, or in-person group meetings) | 24 (30.4) | 34 (43) |
| Personal appointment, one-on-one counseling with a therapist | 17 (20.7) | 38 (46.3) |
| Social worker to connect patients with services (transportation, financial assistance for medical costs) | 31 (38.3) | 34 (42) |
| Coordinator to assist with scheduling medical follow-up appointments | 26 (32.1) | 37 (45.7) |
| Multiple specialty clinic (different doctors located in the same clinic space such as cardiology, pulmonary, psychiatry, etc.) | 13 (16%) | 57 (70.4) |
| Education and providing reliable information (providing information available from trusted sources, e.g., published reputed journals, etc.) | 10 (12) | 57 (68.7) |

### Qualitative analysis of patient’s comments

Overall, 51% of participants responded to the two open ended questions with answers ranging from 3 to 230 words. We categorized comments into the following broad themes: clinic experience, patient health, and “wish list.”

### Clinic experience

Participants addressed their experiences in the PASC clinic and post COVID-19 care received elsewhere. Participants identified scheduling as one area for improvement. However, participants described appreciating when they received quick responses to worries (“*I had an incident… and they got me in … right away*”). Other concerns included distance to the PASC clinic, and the costs of testing and treatment, especially if not covered by insurance. Clinician-patient interaction was an important subtheme. Participants valued open and timely communication. Some participants perceived that some clinicians valued test results more than patients’ perceptions of their health (“*tired of feeling cast off by doctors as just anxiety*”). Participants advised clinicians to listen to their patients, specifically about their symptoms in relation to recommendations for exercise which patients felt were beyond their capabilities. In contrast, participants appreciated when they felt clinicians listened (“*I have never felt so heard and understood*,” “*I was extremely pleased to feel validated with my health and symptoms*,” “*did not treat me like I was just making stuff up*”). Participants understood that knowledge about COVID-19 was evolving, and clinicians were learning along with patients. However, they valued the opportunity to receive treatment and recommendations, including inhalers, pulmonary and physical therapy, and exercise regimens (“*I felt like your clinic actually had an idea after testing of something to do to help*,” “*the treatment I got was spot on, I can get out and do things again*”). They expressed frustration if they felt clinicians had limited suggestions for improvement or treatment.

### Patient Health

Participants often reported ongoing symptoms and frustration with their slow recoveries. They reported that their PASC symptoms such as fatigue could be exacerbated by overexertion or lack of sleep. Participants also volunteered vivid comments describing PASC’s negative impact on their everyday lives and perception of their health before and after COVID-19 infection: “*I understand that there aren’t a lot [of] answers yet. I’m just sick and tired of being sick and tired. Help!*” “*I need to be able to multi-task at [demanding] jobs like I used to have… I’m not sure at all that I could do those jobs as well [now]*.” “*I am sleeping 10-12 hours straight to work full time. I am so tired when I get home*.”

Indeed, for some participants, COVID-19 related health changes and uncertain recovery prospects led to feelings of desperation (“*Feeling pretty hopeless*…,” “*I’m still in pain. I still struggle daily*”). In the words of one participant, “*I feel like a shell of my former self. It’s like being trapped in a not doable stage of life*.”

### Wish List

Participants had recommendations about scheduling, referrals, testing, and treatment. They suggested that speedy initial and follow-up appointments, timely referrals, and consistent follow-up and coordination would improve patient experience and allay frustration with ongoing symptoms. They described valuing continuity of care and wanting their providers to share information. They also sought access to new types of care and specialties, for example, through a multidisciplinary clinic or referrals to a wider range of providers. Overlapping with this interest in specialty care, participants requested additional tests and treatments. Requested avenues for exploration included COVID-19 impact on hormones, and additional cardiac or pulmonary testing, including during exertion. Requested treatments included cognitive and behavioral rehabilitation or therapy, and cardiac rehabilitation. Additional suggestions included support groups for patients and bilingual services.

Research represented a subtheme of interest in this category. Participants wanted to know that clinicians were well-educated on disorders (e.g., dysautonomia) and up to date on current research (e.g., on hormonal disruption) and experimental therapies. One participant suggested clinicians seek information across research and clinical centers about “*what’s worked for them, and things being tried*.” In addition, responses indicate that patients are interested in participating in research and want to hear updates from their clinicians (“*I wish someone would keep us updated with new advancements*,”, “*I hope we will be notified as treatments become a thing for people with lasting symptoms*”).

## Discussion

Our mixed-method survey study highlighted experiences of patients with PASC, the impact of symptoms on their health and their experience with healthcare. The timely feedback by patients for a newly developed PASC clinic in the first year helped us understand barriers and identify key areas of improvement. In our PASC clinic, patients were offered particularly detailed assessment including pulmonary evaluation and behavioral health screening at a single visit. To date various studies have reported multiple symptoms in PASC. ^7-10^ Within our study’s assessment of most concerning PASC symptoms, patients reported fatigue (52%), “brain fog” (39%), and shortness of breath (30%). Our study is in line with previous studies that have reported negative impact on quality of life in COVID-19 survivors. ^7, 8^ On PROMIS measures, patients reported significantly reduced functioning (≥1.5 SD below mean) in their physical health (22.5%), mental health (16%) and cognitive abilities (18%). Interestingly, symptoms during acute illness did not relate to reduced scores, but patients who reported multiple symptoms at three months scored low in physical, mental, and cognitive PROMIS measures. Therefore, we recommend interval follow up of the patients after SARS-CoV-2 infection irrespective of the severity of initial infection. Primary care professionals should consider screening patients with history of SARS-CoV-2 infection for PASC during routine visits.

In our study, approximately 30% of patients reported an increase in mental health symptoms (including anxiety, depression and sleeping difficulties) within three months of infection. A recent metanalysis reported high prevalence of anxiety (1 in 3), sleeping difficulties (1 in 4), depression (1 in 5) and post traumatic disorder (1 in 8) in COVID-19 survivors. ^33^ Multiple other studies are consistent with our findings and emphasize the need to address mental health and sleeping difficulties in patients with PASC. ^2, 5, 34-36^ We integrated behavioral health assessment in our PASC clinic by incorporating anxiety and depression self-reporting questionnaires. If appropriate based on overall assessment and discussion of screen results with the patients, they were also referred to a psychologist. Adopting established practices like incorporating simple distress thermometers can also be used to empower clinicians to provide psychosocial support for patients. ^37^

The qualitative analysis of patient comments adds nuance and context to some of the quantitative results and highlights the profound impact of symptoms on patients’ everyday lives. Our study echoes result from other studies where patient with PASC preferred continuity of care, multidisciplinary rehabilitation, and attentive listening. ^6, 38, 39^ Therefore, we advocate for adequate appointment time for detailed assessment and counselling. ^40^ Multiple co-located specialties in a same clinic may not be possible given multisystemic symptoms which may need several different providers for several concerns in one day. However, in our PASC clinic we established several partnerships with different specialties including behavioral health, neuropsychology (follow up for cognitive assessment) and pulmonology (follow up for chronic lung disease) with identified clinical ‘champions’ who provided subspeciality assessment in a timely manner.

Our experience brings to attention the unique challenges and needs of patients in rural areas with long wait times for appointments, long distances travelled for health care assessment and financial concerns with limited access to healthcare. Further referral to different specialties adds to care complexity along with cost and time lost from work and additional travel. ^14, 16^ Nevertheless, the appointment wait times and distance travelled reported in this study should be contextualized; our large academic center provides resources for patients across the state and operates the only PASC clinic in the area. Partnering with community leaders at outreach clinics (e.g., established UIHC outreach clinic at locations across the state) and local resources (e.g., physical rehabilitation centers) could strengthen the referral system. We also provided follow up care with telemedicine for established patients. In addition, a dedicated clinical nurse coordinator was appointed to assist and coordinate care, especially scheduling tests, subspeciality referrals and follow up care. Furthermore, adequate steps should be taken by policymakers and stakeholders to alleviate the additional economic burden related to evaluation and management of PASC. ^41, 42^

Several limitations of our study warrant consideration. The sample was primarily women, Caucasian, and symptomatic; therefore, the findings are less generalizable to men, different racial/ethnic groups, and asymptomatic patients. We were not able to send the survey in other languages like Spanish which may have decreased survey responses from the Spanish-speaking population. Other races were significantly unrepresented in our survey; however, it is also representative of the demographics of midwestern rural Iowa.^43^ This study is not generalizable to patients in urban areas, or patients seeking care at community and private settings. Patients with confirmed positive SARS-CoV-2 tests were seen in our PASC clinic which excludes patients who were not tested during pandemic’s initial phase. Not all the participants answered the open-ended questions, thus some participant perspectives are not represented. Nevertheless, our analysis of open-ended responses validated and contextualized the survey quantitative results. There is still possibility of recall bias for recollection of dates and symptoms. Finally, there is possibility of recovery of an unknown proportion of patients may have affected survey responses.

## Conclusion

Our study reports high PASC symptom burden, its impact on health and patient experience with healthcare. It is important that primary care professionals listen to patients with PASC with empathy and support them during recovery. Healthcare systems and policymakers should focus on accessible, comprehensive, and patient-centered integrated care.

## Supporting information

The survey (Appendix 1) was drafted using Qualtrics (Qualtrics, Provo, UT)

The odds of expressing reduced physical health, mental health, and cognitive function did not differ by sex or initial care (outpatient vs. inpatient;

## Data Availability

All data produced in the present work are contained in the manuscript

## Appendices

**Appendix 1.**Survey questionnaire.

**Appendix 2.**Odds of expressing reduced Global Physical Health, Global Mental Health, and Cognitive Functioning Abilities by gender and type of care received

## Acknowledgement

The authors would like to acknowledge the staff at University of Iowa Post-COVID-19 clinic who assisted in caring for our patients. The authors also thank Kristina Greiner for her editing assistance.

## Notes

Conflict of Interest: None

Funders: Research reported in this publication was supported by the National Center for Advancing Translational Sciences of the National Institutes of Health (NCATS) under Award Number UL1TR002537

### Competing Interest Statement

The authors have declared no competing interest.

### Funding Statement

Research reported in this publication was supported by the National Center for Advancing Translational Sciences of the National Institutes of Health (NCATS) under Award Number UL1TR002537

### Author Declarations

IRB of University of Iowa hospitals and clinics gave ethical approval for this work

