## Supplementary material for "Patient experience with healthcare: Feedback for a ‘Post COVID-19 clinic’ at a tertiary care center in rural area": The survey (Appendix 1) was drafted using Qualtrics (Qualtrics, Provo, UT)

Post-COVID Clinic Study

**Following Questions are related to COVID infection and your health**

Q 1. Time of positive COVID test (approximately):

Date MM/DD/YYYY ________________________________________________

Q 2. Level of medical care received during your initial COVID-19 infection? (Select all that apply)

•	Managed at home without medical care

•	Telemedicine visits only

•	Urgent care visit

•	Emergency room visit and discharged

•	Admitted to the hospital

•	Admitted and cared for in an intensive care unit (ICU)

•	Other ________________________________________________

Q 3-8. Symptoms at time COVID Infection (choose all that apply) and Symptoms (> 3 months) after COVID infection

| Body systems | Symptoms | During initial COVID illness when first tested | After COVID (> 3 months after COVID test) |
| --- | --- | --- | --- |
| 1. General | Fatigue |  |  |
|  | Fever |  |  |
|  | Exercise intolerance |  |  |
|  | Hair loss |  |  |
|  | New pain |  |  |
| 2. Neurological/Nerves/Memory | Headache |  |  |
|  | Brain fog (memory and/or other cognitive problems) |  |  |
|  | Loss of smell |  |  |
|  | Loss of taste |  |  |
|  | Numbness |  |  |
|  | Tingling/pins and needles in extremities |  |  |
|  | Fainting/blackout |  |  |
|  | Tremors/shakiness |  |  |
|  | Ringing in the ears |  |  |
| 3. Lung | Cough |  |  |
|  | Shortness of breath |  |  |
|  | Pain on breathing |  |  |
| 4. Heart | Chest pain |  |  |
|  | palpitations |  |  |
|  | Feeling lightheaded |  |  |
|  | Leg swelling |  |  |
| 5. Gastrointestinal | Nausea |  |  |
|  | Vomiting |  |  |
|  | Abdominal pain |  |  |
|  | Diarrhea |  |  |
|  | Constipation |  |  |
|  | Loss of appetite |  |  |
| 6. Mental health | Anxiety |  |  |
|  | Depression |  |  |
|  | Sleep problems |  |  |

Q 9. Please share the three symptoms that you are most concerned about **right now**.

•	1st ________________________________________________

•	2nd ________________________________________________

•	3rd ________________________________________________

•	Something else? (Please share) ________________________________________________

Q 10. Please answer the following questions about your overall health AFTER COVID Infection

|  | Poor | Fair | Good | Very good | Excellent |
| --- | --- | --- | --- | --- | --- |
| In general, would you say your health is: |  |  |  |  |  |
| In general, would you say your quality of life is: |  |  |  |  |  |
| In general, how would you rate your physical health? |  |  |  |  |  |
| In general, how would you rate your mental health, including your mood and your ability to think? |  |  |  |  |  |
| In general, how would you rate your satisfaction with your social activities and relationships? |  |  |  |  |  |

Q 11. To what extent are you able to carry out your everyday physical activities such as walking, climbing stairs, carrying groceries, or moving a chair?

•	Not at all

•	A little

•	Moderately

•	Mostly

•	Completely

Q 12. In the past 7 days How would you rate your pain on average? (0 - No pain) -(10- Worst imaginable pain)

Q 13. In the past 7 days How would you rate your fatigue on average?

•	None

•	Mild

•	Moderately

•	Severe

•	Very Severe

Q 14. In general, please rate how well you carry out your usual social activities and roles. (This includes activities at home, at work and in your community, and responsibilities as a parent, child, spouse, employee, friend, etc.)

•	Poor

•	Fair

•	Good

•	Very Good

•	Excellent

Q 15. In the past 7 days How often have you been bothered by emotional problems such as feeling anxious, depressed, or irritable?

•	Never

•	Rarely

•	Sometimes

•	Often

•	Always

Q 16. Please respond to each question or statement by marking one box per row

|  | Not at all | A little bit | Somewhat | Quite a bit | Very much |
| --- | --- | --- | --- | --- | --- |
| My mind has been sharp as usual |  |  |  |  |  |
| My Memory has been as good as usual |  |  |  |  |  |
| My thinking has been as fast as usual |  |  |  |  |  |
| I have been able to keep track of what I am doing, even if I am interrupted |  |  |  |  |  |

**The next questions are about your experience with the Respiratory Illness Follow-up Clinic (i.e., Post-COVID Clinic) at the University of Iowa**

Q17. How much time does it take you to travel to the Post-COVID clinic (one way)?

•	Under 30 minutes

•	30 minutes - 1 hour

•	1 hour - 2 hours

•	2 hours - 3 hours

•	3 hours - 4 hours

•	4 hours - 5 hours

•	More than 5 hours

Q 18. How long did you have to wait for your first appointment at the Post-COVID Clinic?

•	Less than 2 weeks

•	2 weeks to 1 month

•	1 month to 2 months

•	2 months to 3 months

•	3 months to 4 months

•	over 4 months

Q 19. How satisfied were you with the following aspects of the Post-COVID Clinic at University of Iowa?

|  | Very dissatisfied | Dissatisfied | Neutral | Satisfied | Very satisfied | Not applicable |
| --- | --- | --- | --- | --- | --- | --- |
| Scheduling clinic appointments |  |  |  |  |  |  |
| Getting lung function tests (also known as PFTs or spirometry) |  |  |  |  |  |  |
| Getting CT scan |  |  |  |  |  |  |
| Getting blood tests |  |  |  |  |  |  |
| Interaction with the front desk staff and Medical assistant |  |  |  |  |  |  |
| Interaction with the doctor in the Post-COVID Clinic |  |  |  |  |  |  |
| Scheduling referrals / care coordination for subspecialities like cardiology, psychiatry, rheumatology, rehabilitation medicine (OT or PT) |  |  |  |  |  |  |
| Overall, addressing your physical health concerns |  |  |  |  |  |  |
| Overall, addressing your mental health |  |  |  |  |  |  |

Q 20. Please check all the specialists that were recommended by the doctor (including your primary care doctor) for evaluation of your post-COVID health concerns

•	Cardiology (Heart specialist)

•	Rheumatology (Joint specialist)

•	Neurology (Nervous system specialist)

•	Psychiatry (Mental health Specialist)

•	Neuropsychology (Psychologist)

•	Pulmonary (Lung specialist)

•	Ear, Nose, Throat Doctor (ENT)

•	Gastroenterologist

•	Physical therapy/ Occupational therapy

•	Pulmonary rehabilitation

•	Other _______________________________________________

Q 21. Which referrals have you scheduled to complete or already completed?

•	Cardiology (Heart specialist)

•	Rheumatology (Joint specialist)

•	Neurology (Nervous system specialist)

•	Psychiatry (Mental health Specialist)

•	Neuropsychology (Psychologist)

•	Pulmonary (Lung specialist)

•	Ear, Nose, Throat Doctor (ENT)

•	Gastroenterologist

•	Physical therapy/ Occupational therapy

•	Pulmonary rehabilitation

•	Other _______________________________________________

Q 22. Which of the following are barriers to care/reason for not being able to attend?

•	Delay in appointment

•	Could not get appointment

•	Travel long distance to come for appointments

•	Complex care-Multiple clinic visits with different providers

•	Limited by symptoms so unable to attend

•	Unable to miss work to attend appointments

•	Prefer Telemedicine appointment than in-person due to COVID risk

•	Financial concerns/don’t know if insurance will cover tests and appointments

•	Any other barriers, please feel free to add ________________________________________________

Q 23. How helpful do you think each of the following resources could be to you or other patients with post-COVID symptoms?

|  | Not at all helpful | Slightly helpful | Somewhat helpful | Very helpful | Extremely helpful |
| --- | --- | --- | --- | --- | --- |
| **Continuity of Care** and regular follow up with same doctors |  |  |  |  |  |
| **Physical Therapy/Occupational Therapy** (rehabilitation to increase physical strength and exercise capacity) |  |  |  |  |  |
| **Pulmonary Rehabilitation** Rehabilitation to improve breathing (supervised program) |  |  |  |  |  |
| **Informal support group** open forum e.g., Online support group with other patients with similar experiences |  |  |  |  |  |
| **Group therapy** intervention led by a mental health provider eg on zoom or in-person group meetings |  |  |  |  |  |
| **Personal Appointment** One on one counseling with a therapist |  |  |  |  |  |
| **Social worker** to connect patients with services (transportation, financial assistance for medical costs) |  |  |  |  |  |
| **Coordinator** to assist with scheduling medical follow-up appointments |  |  |  |  |  |
| **Multiple specialty clinic** (different doctors located in the same clinic space such as cardiology, pulmonary, psychiatry etc.) |  |  |  |  |  |
| **Education and providing reliable information** (Providing information available from trusted sources e.g., published reputed journals etc.) in emails or frequent updates |  |  |  |  |  |

Q 24. Please identify any other resources you think would be helpful in an ideal post-COVID Clinic:

________________________________________________________________

________________________________________________________________

________________________________________________________________

________________________________________________________________

________________________________________________________________

Q 25. Is there anything else you would like to share with us? Please feel free to give us any feedback:

________________________________________________________________

________________________________________________________________

________________________________________________________________

________________________________________________________________

________________________________________________________________

**Please answer the following questions about yourself**

Q 26. How old are you?

•	18-29

•	30-39

•	40-49

•	50-59

•	60-69

•	70 or over

Q 27. Gender (Select all that apply)

•	Man

•	Woman

•	Non-binary

•	Prefer not to say

•	Prefer to self-describe ________________________________________________

Q 28. Ethnicity (Select all that apply)

•	Asian

•	American Indian or Pacific Islander

•	African American

•	Caucasian

•	Latino/ Hispanic

•	Two or more races

•	Prefer not to answer

•	Prefer to self-describe ________________________________________________
