## Supplementary material for "Patient experience with healthcare: Feedback for a ‘Post COVID-19 clinic’ at a tertiary care center in rural area": The odds of expressing reduced physical health, mental health, and cognitive function did not differ by sex or initial care (outpatient vs. inpatient;

| **Appendix 2.** Odds of Expressing Reduced Global Physical Health, Global Mental Health, and Cognitive Functioning Abilities by Gender and Type of Care Received | | | | | |
| --- | --- | --- | --- | --- | --- |
| **Variable** | | **Physical Health Frequency (%)** | | **OR (95% CI)** | ***p*-Value^1^** |
|  |  | **Mean±1.49SD** | **Reduced ≥ 1.5 SD** |  |  |
| Sex | |  |  |  |  |
|  | Male | 19 (76%) | 6 (24%) | 0.89 (0.29, 2.78) | 0.845 |
|  | Female | 39 (78%) | 11 (22%) |  |  |
| Outpatient | |  |  |  |  |
|  | No | 10 (58.8%) | 7 (41.2%) | 0.3 (0.09, 0.97) | 0.038 |
|  | Yes | 48 (82.8%) | 10 (17.2%) |  |  |
| Inpatient | |  |  |  |  |
|  | No | 45 (80.4%) | 11 (19.6%) | 1.89 (0.59, 6.09) | 0.345 |
|  | Yes | 13 (68.4%) | 6 (31.6%) |  |  |
|  | | **Mental Health Frequency (%)** | |  |  |
|  |  | **Mean±1.49SD** | **Reduced ≥ 1.5 SD** |  |  |
| Sex | |  |  |  |  |
|  | Male | 23 (76.7%) | 7 (23.3%) | 0.45 (0.14, 1.44) | 0.171 |
|  | Female | 51 (87.9%) | 7 (12.1%) |  |  |
| Outpatient | |  |  |  |  |
|  | No | 16 (84.2%) | 3 (15.8%) | 1.01 (0.25, 4.07) | 1.000 |
|  | Yes | 58 (84.1%) | 11 (15.9%) |  |  |
| Inpatient | |  |  |  |  |
|  | No | 57 (83.8%) | 11 (16.2%) | 0.91 (0.23, 3.66) | 1.000 |
|  | Yes | 17 (85%) | 3 (15%) |  |  |
|  | | **Cognitive Function Abilities Frequency (%)** | |  |  |
|  |  | **Mean±1.49SD** | **Reduced ≥ 1.5 SD** |  |  |
| Sex | |  |  |  |  |
|  | Male | 24 (77.4%) | 7 (22.6%) | 0.61 (0.2, 1.82) | 0.368 |
|  | Female | 51 (85%) | 9 (15%) |  |  |
| Outpatient | |  |  |  |  |
|  | No | 15 (78.9%) | 4 (21.1%) | 0.75 (0.21, 2.66) | 0.736 |
|  | Yes | 60 (83.3%) | 12 (16.7%) |  |  |
| Inpatient | |  |  |  |  |
|  | No | 58 (81.7%) | 13 (18.3%) | 0.79 (0.2, 3.09) | 1.000 |
|  | Yes | 17 (85%) | 3 (15%) |  |  |
| ^1^Groups compared using chi-square tests of independence and Fisher’s exact test, as appropriate. | | | | | |
